# Predicting Graduation in Undergraduate Medical Education: A Machine Learning Analysis Across Diverse High School Curricula

**DOI:** 10.64898/2026.03.07.26347831

**Authors:** Jalal Mohamadeya, Amar Khamis, Laila Alsuwaidi, Aida Joseph Azar

**Affiliations:** Digital and AI Department, Mohammed Bin Rashid University of Medicine and Health Sciences, Dubai Health, Dubai, United Arab Emirates; Hamdan Bin Mohammed College of Dental Medicine, Mohammed Bin Rashid University of Medicine and Health Sciences, Dubai Health, Dubai, United Arab Emirates; College of Medicine, Mohammed Bin Rashid University of Medicine and Health Sciences, Dubai Health, Dubai, United Arab Emirates; Student Affairs, Mohammed Bin Rashid University of Medicine and Health Sciences, Dubai Health, Dubai, United Arab Emirates

**Keywords:** Machine Learning, Prediction, Doctor of Medicine, Undergraduate, Higher Education

## Abstract

**Background:** The United Arab Emirates (UAE) is characterised by a diverse educational landscape, where students enter medical school from various high school curricula. Understanding how these varied academic backgrounds influence medical students’ academic performance is essential. The transition to medical school is a critical phase, with graduation outcomes carrying important implications for both students and institutions. Identifying early predictors of success is crucial to improving student support and academic outcomes in undergraduate medical education.

**Aim:** This study aimed to evaluate the predictive value of high school curriculum type on graduation outcomes in an undergraduate medical education program.

**Methods:** A retrospective cohort study was conducted on undergraduate medical students enrolled at Mohammed Bin Rashid University of Medicine and Health Sciences (MBRU), Dubai Health, Dubai, UAE, from its inception in 2016 through 2024. The data were accessed for this research on 04/06/2024. The study employed machine learning methods, including Bayesian Networks (BN), Neural Networks (NN), and Random Forests (RF), to evaluate the predictive power of high school curriculum type and other academic variables for graduation success.

**Results:** The study included 661 undergraduate medical students, predominantly female, 76.7% (n=507). Students represented 11 high school curricula, with the American (48.1%) and British (22.7%) systems being the most common. Among 122 students eligible to graduate, the Bayesian Network model demonstrated the highest predictive accuracy (AUC = 0.94). The cumulative GPA was the most influential predictor. The model correctly identified 269 out of 494 students (54.5%) as likely to graduate.

**Conclusion:** The type of high school curriculum alone is not a strong predictor of graduation success. Academic performance during medical school and providing targeted support for students from diverse educational backgrounds are more robust predictors. Advanced predictive modelling holds promise for educational research and institutional policy development.

## INTRODUCTION

Extensive research underscores a strong correlation between the rigor of secondary school education and subsequent academic success in higher education. Students who engage in rigorous academic programs, such as Advanced Placement (AP) or the International Baccalaureate (IB), tend to perform better in university, achieving higher grade point averages and improved graduation rates (Al-Mazrou AM, 2008). To predict academic outcomes, various modelling techniques, including logistic regression, decision trees, and neural networks, have been employed, incorporating factors such as secondary school performance, standardized test scores, and socio-economic background (Maslov Kruzicevic et al., 2012).

In medical education, studies indicate that the quality of secondary education and curriculum type have been shown to significantly influence performance on key assessments such as the Medical College Admission Test (MCAT), a standardized exam for medical school entry, and the United States Medical Licensing Examination (USMLE) a multi-step exam required for medical licensure in the U.S. (Chan LJ et al., 2022). More recently, machine learning approaches have also proven useful in forecasting student outcomes, assisting institutions in refining retention strategies, and optimizing resource distribution (Hammoudi Halat et al., 2023). Data from various global contexts further support the role of secondary education, admission policies, and standardized testing in shaping success in medical and health sciences programs (Tamimi, A. et al., 2023; Qahmash A et al., 2023).

As the global demand for competent healthcare professionals grows, medical schools are increasingly tasked with identifying candidates most likely to succeed in academically rigorous undergraduate medical programs (Frenk J et al., 2010). A key component of the medical school selection process is the type of secondary school curriculum. While standardized tests and grade point averages (GPA) are commonly used admission criteria, the extent to which different academic systems (e.g., national curricula, IB, British A-levels, and American high school diplomas) predict long-term success in medical education remains insufficiently explored.

Dubai, in the United Arab Emirates (UAE), offers a unique setting for such an investigation. With more than 17 curricula represented across its private schools and a highly diverse student population, Dubai provides a valuable context for analysing how pre-university academic background influences medical school performance (KHDA, 2023 https://web.khda.gov.ae/en/resources/khda%E2%80%99s-data-statistics). Although previous studies have mainly focused on isolated metrics such as GPA, standardized test results, and aptitude assessments, few have considered the broader pedagogical and contextual differences among secondary education systems (Kreiter CD et al., 2013; K. Monroe et al., 2013; Patterson F et al., 2016). These curricula vary not only in context and evaluation methods but also in their emphasis on critical thinking, problem-solving, and independent learning, skills essential for success in medical training. Recent research by Tamimi et al. (2023) further underscores this point, demonstrating that students from different high school curricula, such as the IB and British systems, perform differently in medical school, highlighting the influence of educational background on academic outcomes (*Tamimi, A., Hassuneh, M., Tamimi, I. et al. Admission criteria and academic performance in medical school. BMC Med Educ 23, 273 (2023)*. https://doi.org/10.1186/s12909-023-04251-y).

Selection criteria often prioritize standardized test scores and overall academic performance, however, there is limited empirical evidence linking specific secondary school curricula to long-term success in medical education (Patterson F et al., 2016). Understanding how diverse academic backgrounds influence program completion and academic outcomes can help undergraduate medical institutions refine their selection processes to admit candidates with the highest probability of success. Moreover, this will offer valuable guidance to students, parents, and educators on the strengths and limitations of various curricula in preparing individuals for medical studies. These insights contribute to the development of data-driven admission policies and targeted academic support frameworks, ultimately enhancing the quality and effectiveness of medical education in the UAE.

This study aimed to address this gap by examining the relationship between secondary school curricula and graduation outcomes in undergraduate medical education, with the goal of informing student selection policies and academic support strategies within the UAE’s academically diverse education landscape.

## MATERIALS AND METHODS

### Study Setting

This retrospective cohort study was conducted at Mohammed Bin Rashid University of Medicine and Health Sciences (MBRU), a medical university operating under Dubai Health in Dubai, United Arab Emirates. Although the program confers a primary medical qualification, it is referred to generically here to avoid specifying degree titles such as Doctor of Medicine (MD) or Bachelor of Medicine, Bachelor of Surgery (MBBS). Students are eligible for admission to the College of Medicine (CoM) upon completion of their secondary education and fulfillment of the admission criteria for the medical school. Established in 2016, the study focused on undergraduate students enrolled in the medical degree program, using institutional data collected from the university’s inception through 2024.

### Study Population

All students enrolled in the CoM at MBRU from the university’s inception in 2016 through 2024 were considered eligible for inclusion in this study. June 2024 marks the year the study was initiated, and the most recent cohort data became available. A comprehensive dataset was compiled for applicants and students who applied to the CoM. Each record included detailed information such as applicant and student IDs, the year of admission, personal and demographic data (including gender and country of residence), academic background, interview performance assessed through Multiple Mini Interviews (MMI), high school curriculum details, and English language proficiency. For those who enrolled in the medical program during this period, additional layers of student-specific information were collected. These records featured a unique student identifier, updated academic profiles, course registration, transcript data, and enrolment status (categorized as active, enrolled, withdrawn, or graduated). Student academic performance was assessed using the most recent cumulative GPA, which reflects cumulative academic achievement across the program. Further, course-level data were extracted from the university’s Learning Management System (LMS) to enrich the dataset. This included information on course enrolments, coursework records, and final course grades, which were reported using either a letter grade system (ranging from A to F) or a Pass/Fail scheme. Student records were consolidated by integrating data from both the admissions system and the LMS. These finalized datasets were organized and stored in spreadsheet format to facilitate subsequent statistical analysis.

*Figure 1* illustrates the stages of the applicant-to-student journey and outlines the data collection process across 6,784 applications submitted to the MD program between 2016 and 2024. From these, 661 applicants were successfully admitted and enrolled, forming the core dataset analysed in this study.

**Figure 1.**
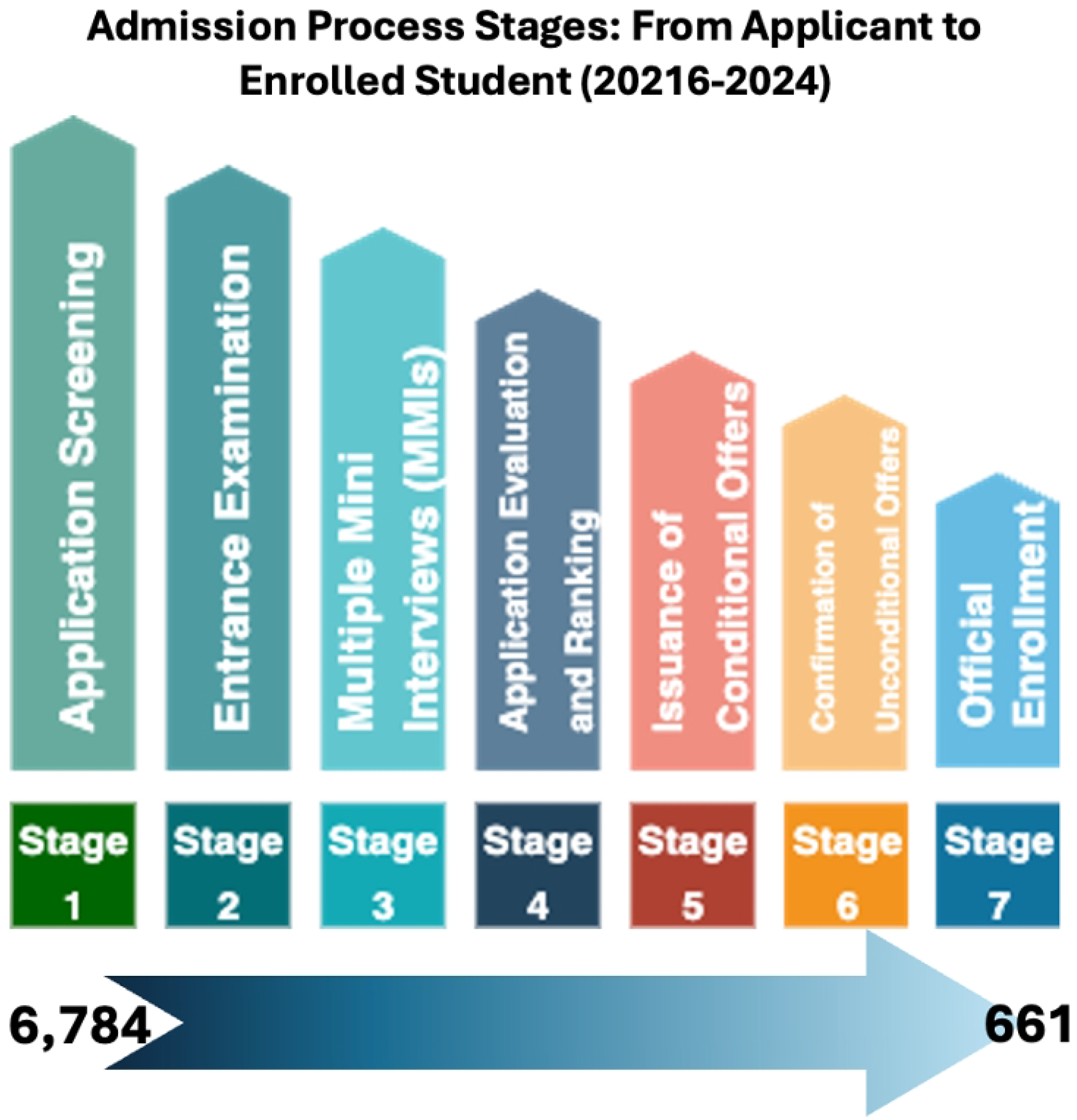

### Design and Techniques

The collected data for the students who enrolled in the CoM from its inception in 2016 and 2024 were divided into two datasets for analysis. The first, termed the training file, included data from 167 students who enrolled between 2016 and 2018, had completed their studies, and graduated by the time of analysis; this dataset was used to train predictive models. The second dataset, referred to as the prediction file, contained data for the students who enrolled between 2019 and 2024 and had not yet graduated. Machine Learning (ML) techniques were applied to the training data to identify patterns. These patterns were then used to forecast academic outcomes for the prediction dataset using two different scenarios within an auto-classifier model, which automatically categorizes students based on their likelihood of graduation.

### Prediction Models

IBM SPSS Modeler (IBM North America, New York, USA) is a data science and machine learning platform that facilitates predictive modelling without the need for complex coding. It supports the entire modelling process, from data cleaning and preparation to algorithm selection and model deployment. In this study, SPSS Modeler was used to predict the graduation status of a new cohort of students. By utilizing built-in algorithms and visualization tools, the software identified patterns and relationships within the data to estimate each student’s likelihood of graduating.

The software explored 14 potential predictive models in its Auto-classifier mode, including C5.0 Decision Tree Algorithm (C5), Logistic Regression (LR), Bayesian Network (BN), Linear Discriminant Analysis (D), Linear Support Vector Machine (LSVM), Random Tree (RT), Extreme Gradient Boost Linear (XGBL), Extreme Gradient Boost Tree (XGBT), Chi-Square Automatic Interaction Detection (CHAID), Quick Unbiased Efficient Statistical Tree (QUEST), Classification and Regression Tree (C&R Tree), Neural Network (NN), Decision List (DL), and Tree AS. It automatically selects the top 5-6 models and provides the results sequentially. In the training dataset, we used the following 8 independent variables associated with graduation status as predictors: gender, curriculum, country, year of admission, MMI score, semester GPA, year GPA, and cumulative GPA. After extensive training, the model was ready to predict the graduation status of the new cohort of students in the dataset. Graduation status for the cohorts admitted during the academic years 2016-2017, 2017-2018, and 2018-2019 was used as a target variable and defined as a dichotomous variable (graduated or not graduated).

### Validation and Testing

Python code was developed to validate the results produced by IBM SPSS Modeler. Model performance was evaluated using cross-validation and holdout test sets, with key metrics including area under the curve (AUC), accuracy, sensitivity, and specificity. While internal validation was completed, external validation should be performed if multicenter data is available.

### Statistical Analysis

SPSS for Windows, version 29.0 (SPSS Inc., Chicago, IL), was used to perform descriptive statistical analysis of the high school curriculum variables. This analysis summarised key features of the dataset, including frequency counts, percentages, standard deviations, and ranges, to provide an overview of the distribution and diversity of students’ educational backgrounds.

### Data Security and Confidentiality

Measures were implemented to ensure the security and confidentiality of student data in compliance with ethical and legal standards. All data were anonymized, with access restricted to authorized personnel only. Data were encrypted during storage and transmission and stored as password-protected soft copies. Student anonymity was rigorously preserved throughout the study.

### Ethical Approval

The study was approved by the relevant MBRU IRB Committee (Reference # MBRU IRB-2024-205) and by the Dubai Scientific Research Ethics Committee (DSREC), Dubai Health Authority (DSREC-GL10-2024).

## RESULT

### Demographics

*Table 1* presents the demographic characteristics of the study sample, which comprised 661 undergraduate medical students enrolled at MBRU from its establishment in 2016 to 2024. The majority of students were female, 507 (76.7%), while males accounted for 154 (23.3%). Regarding institutional classification, most students, 573 (94.4%), graduated from private schools, with only 34 (5.6%)from government schools. The distribution of students by academic year among the 661 participants shows a clear upward trend over time. In 2016, the year MBRU was established, student representation was at 8.6%, followed by a slight decline to 7.6% in 2017, the lowest point during the observed period. From 2018 onward, the data revealed a gradual and consistent increase, reaching 12.3% in 2023. The most notable shift occurred in 2024, when there was a substantial spike to 18.6%, marking the highest annual intake.

**Table 1:** Demographic and Educational Characteristics of Undergraduate Medical Students at the CoM, MBRU, UAE (2016 to 2024, N = 661) No. Number; CoM, College of Medicine; MBRU, Mohammed Bin Rashid University of Medicine and Health Sciences, Dubai Health, Dubai, UAE, United Arab Emirates. UAE, United Arab Emirates; SABIS, a global private school network offering an international curriculum.

| Characteristic | No (%) |
| --- | --- |
| **Gender** |  |
| Male | 154 (23.3) |
| Female | 507 (76.7) |
| **High School Location** |  |
| UAE Based | 565 (93.8) |
| Non-UAE Based | 31 (6.2) |
| **Institutional classification** |  |
| Government | 34 (5.6) |
| Private | 573 (94.4) |

Participants also reported a wide variety of curriculum backgrounds. *Figure 2* illustrates the distribution of students by curriculum type. The American Diploma was the most common, comprising 48.1% of the cohort. Following this, the British Curriculum takes the second-largest share, representing 22.7% of the students. This aligns with the widespread popularity of the British education system, renowned for its academic rigor and international recognition. Together with the International Baccalaureate (IB) at 11.5%, these two curricula make up a significant portion, with a combined total of 34.2%. This highlights the strong presence of internationally oriented educational frameworks within the group. However, the distribution also reveals a variety of smaller yet notable educational systems. The Canadian Diploma and the French Baccalaureate each contribute a modest 0.8% to the total, reflecting their relatively limited but still significant representation in the curriculum. Similarly, other national curricula, including the Iranian, Nigerian, and Yemeni systems, each account for less than 1%, which may indicate smaller, more specific communities of students following these paths. Other unique systems, such as the SABIS network (3.6%) and the UAE Government School curriculum (5.6%), also contribute to the distribution.

**Figure 2.**
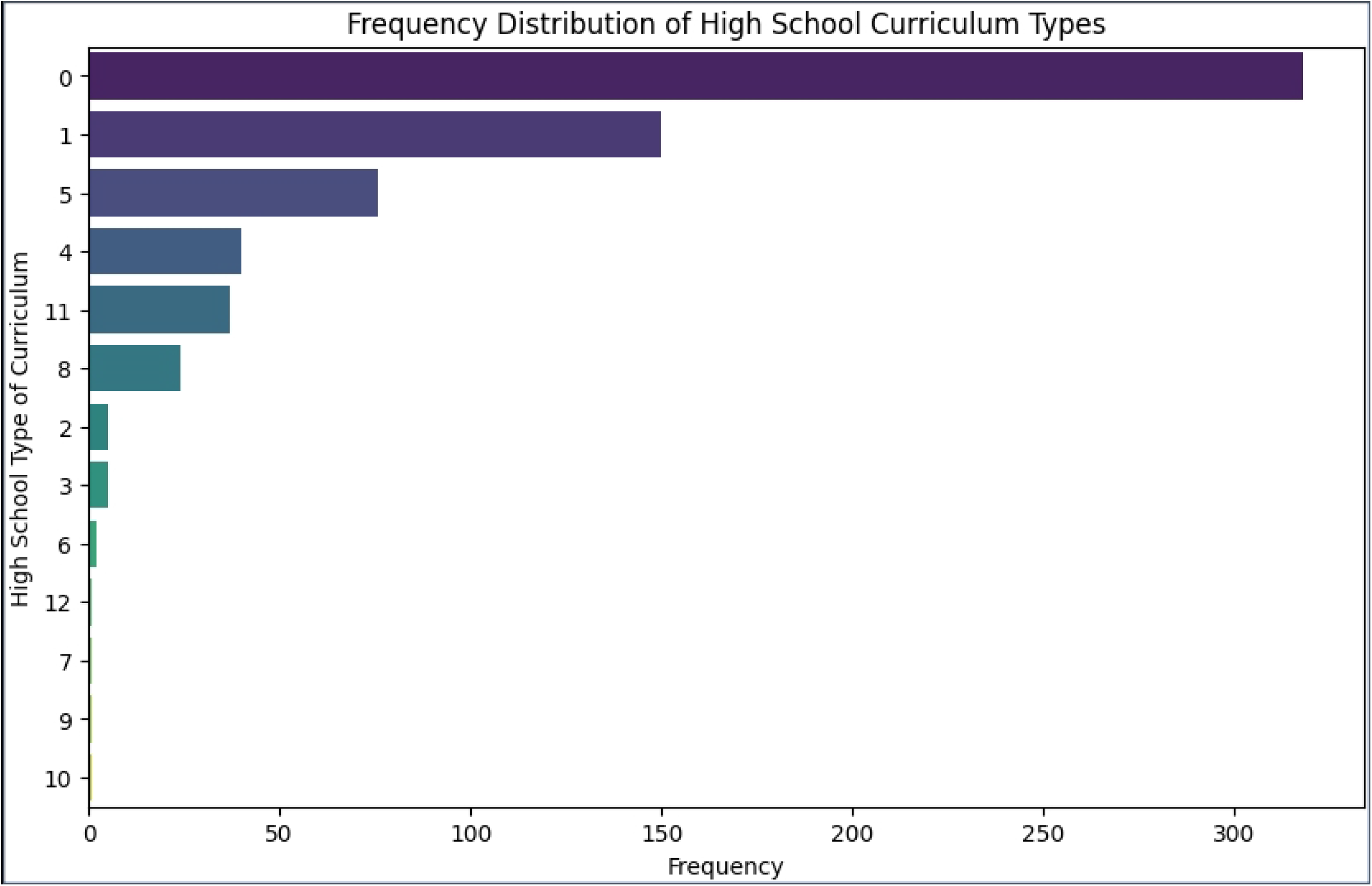
The bar chart illustrates the distribution of undergraduate medical students (N = 661) enrolled at MBRU from 2016 to 2024, based on their high school curriculum background. The most common curricula were the American Diploma (48.1%) and British Curriculum (22.7%). Less common backgrounds included International Baccalaureate, Indian, UAE Government, and SABIS curricula, with several other international curricula represented in smaller proportions.

### Model performance

Using IBM SPSS Modeler software, the target variable was defined as a binary flag indicating whether a student graduated or not, representing the student’s graduation status. The analysis focused exclusively on students enrolled in the Doctor of Medicine program who were eligible to graduate, including 122 students who had already graduated.

*Table 2* presents the performance of five classification algorithms employed to predict graduation outcomes: Bayesian Network (BN), Neural Network (NN), Chi-squared Automatic Interaction Detection (CHAID), Logistic Regression (LR), and Discriminant Analysis (D). Among these models, the Bayesian Network demonstrated the highest predictive performance.

**Table 2:** Quality of Predictor Model: Scenario 2. BN, Bayesian Network; NN, Neural Network; LR, Logistic Regression; DM, Deterministic Model; No., Number; TP, True Positive; TN, True Negative; FP, False Positive; FN, False Negative; SD, Standard Deviation; AUC, Area Under the Curve; Sens, sensitivity; Spec, specificity; PPV, Positive Predictive Value; NPV, Negative Predictive Value.

| Models | No. | Profit (%) | No, field | TP | TN | FP | FN | Accuracy (%) | SD | AUC | Sens. | Spec. | PPV | NPV |
| --- | --- | --- | --- | --- | --- | --- | --- | --- | --- | --- | --- | --- | --- | --- |
| BN | 167 | 80 | 7 | 120 | 32 | 13 | 2 | 91.018 | 0.29 | 0.94 | 0.98 | 0.71 | 0.90 | 0.94 |
| NN | 167 | 81 | 6 | 119 | 33 | 12 | 3 | 91.018 | 0.29 | 0.934 | 0.98 | 0.73 | 0.91 | 0.92 |
| CHAID | 167 | 79 | 3 | 121 | 33 | 12 | 1 | 92.216 | 0.27 | 0.92 | 0.99 | 0.73 | 0.91 | 0.97 |
| LR | 167 | 79 | 8 | 116 | 33 | 12 | 6 | 89.222 | 0.31 | 0.94 | 0.95 | 0.73 | 0.91 | 0.85 |
| DM | 167 | 81 | 5 | 118 | 31 | 14 | 4 | 89.222 | 0.31 | 0.932 | 0.97 | 0.69 | 0.89 | 0.89 |

Cumulative GPA emerged as the most significant predictor of graduation status, while the other variables were ranked equally in terms of importance, as illustrated in *Figure 3*. When the ensemble node was applied to the prediction dataset, the model identified 269 out of 494 students (54.5%) as likely to graduate and 225 students (45.5%) as unlikely to graduate.

**Figure 3.**
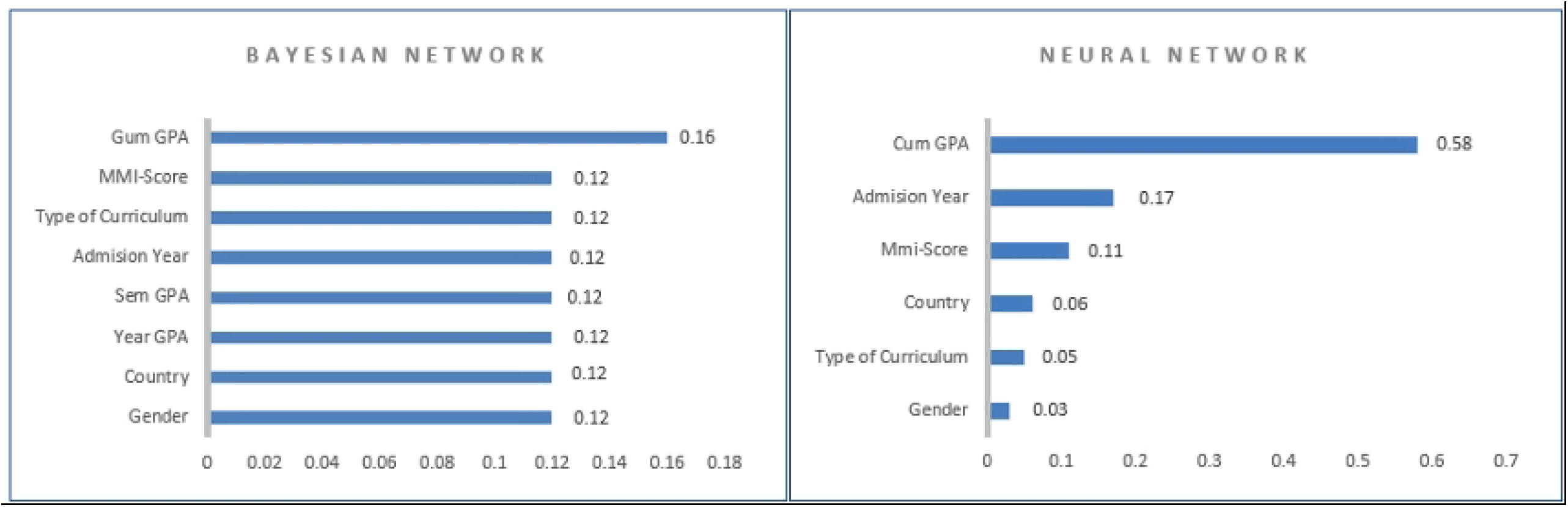
The bar charts show the relative importance of input variables in predicting student graduation status using two models: Bayesian Network (left) and Neural Network (right). In both models, cumulative GPA (Cum GPA) was the strongest predictor. The Bayesian Network assigned equal importance to most other variables, including MMI score, type of curriculum, and admission year. In contrast, the Neural Network assigned varying weights to the remaining predictors, with admission year and MMI score contributing more than factors like gender or type of curriculum.

The ROC curve in *Figure 3* illustrates the model’s performance, with the Bayesian Network (BN) and Logistic Regression (LR) both achieving an AUC of 0.94, while the CHAID model followed closely with an AUC of 0.92.

Feature importance analysis consistently identified cumulative GPA as the most influential factor across models, reinforcing what has been previously suggested in the literature. Semester-wise and annual GPA also emerged as strong predictors. By contrast, gender, high school curriculum, and MMI scores appeared less predictive, though they still contributed marginally to overall model accuracy.

### Model Validity

To ensure the reliability of the predictive models developed in this study, a combination of internal validation techniques and cross-platform verification was employed. The training dataset consisted of medical students from the 2016 to 2018 cohorts, while the prediction models were tested on data from the 2019 to 2024 cohorts. This chronological division was designed to simulate a realistic predictive scenario in which historical data informs future outcomes.

Model performance was primarily assessed using the Area Under the Curve (AUC), along with other metrics, including accuracy, sensitivity, specificity, and predictive values (PPV and NPV). Among all models tested, the Bayesian Network (BN) consistently demonstrated the highest predictive validity, achieving an AUC of 0.94. Logistic Regression and CHAID models also performed robustly, each recording AUC values above 0.92, as shown in *Figure 4*. These results indicate that the models were well-calibrated to distinguish between students who were likely to graduate and those who were not likely to graduate.

**Figure 4.**
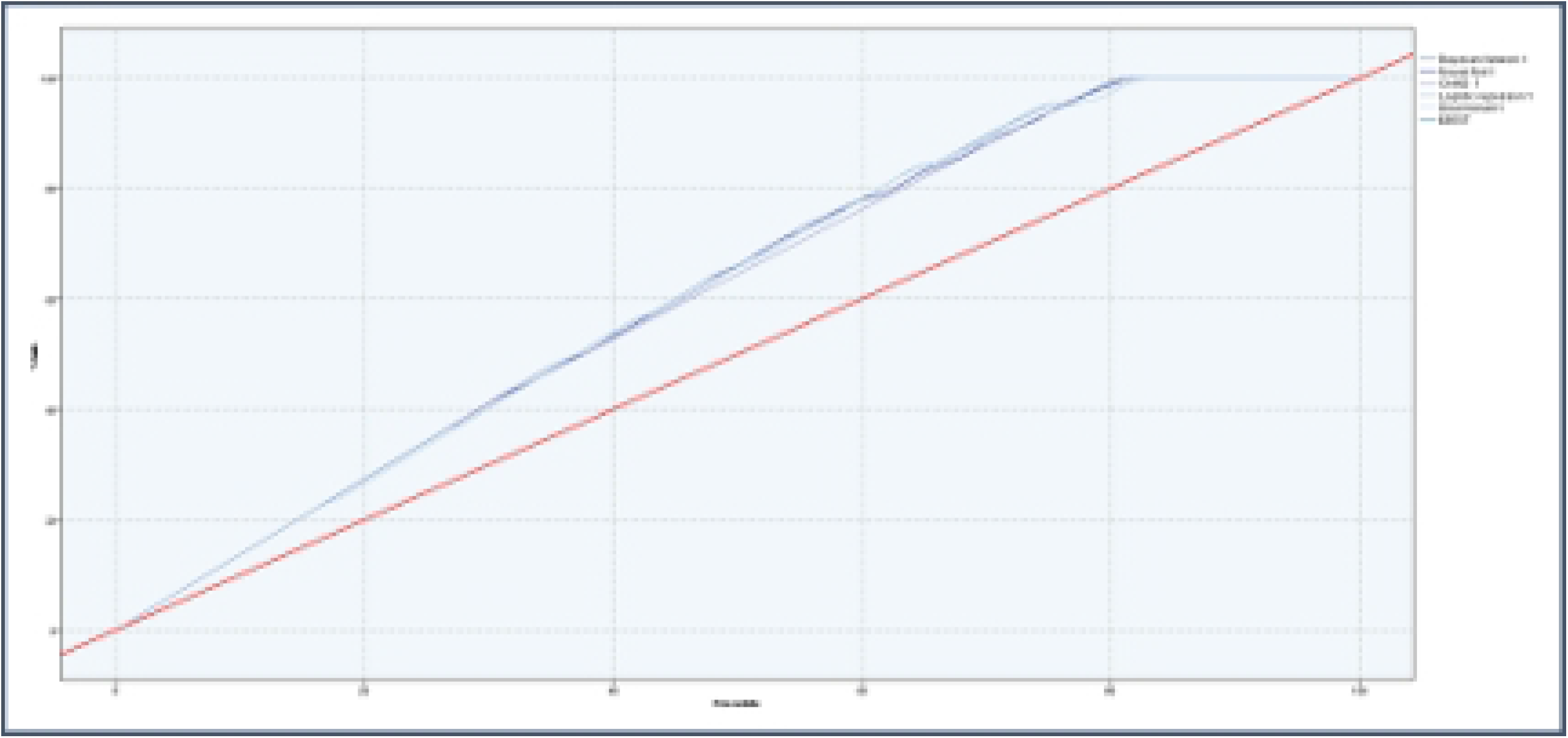

To enhance the credibility of the findings, the analysis was replicated using Python to cross-verify the outcomes generated from SPSS Modeler. This approach enhanced reproducibility and minimized tool-specific biases. While the internal validation results are promising, we recognize the need for future external validation using larger or multi-institutional datasets to confirm the model’s generalizability beyond MBRU.

## DISCUSSION

### Academic prediction of graduation

This study aimed to explore whether a student’s high school curriculum could predict their successful graduation from a medical university, where students are admitted directly after high school into an undergraduate medical program. Using historical institutional data and supervised machine learning techniques, we analyzed various academic and demographic variables to build predictive models of graduation success. The most consistent and important finding across all models was the dominance of cumulative GPA, which reflects overall academic performance, as the strongest predictor of graduation. This aligns with global trends and existing educational literature that emphasize academic performance during university as a key determinant of success (Stegers-Jager et al., 2012; O’Neill et al., 2011).

High school curriculum type was included in the analysis based on previous findings suggesting its potential predictive value in medical education (Yates & James, 2007; Al-Alwan et al., 2023). However, its predictive power was relatively weak compared to cumulative GPA, indicating that students’ performance during the medical program is a more accurate predictor of their likelihood of graduation.

### Curriculum Diversity

Notably, while American and British high school curricula were most common among the sample, students from over ten different educational systems were represented. This level of diversity reflects the multicultural and international context of medical education in the UAE, consistent with findings from other medical schools that serve globally mobile student populations (Chen et al., 2012; Harden, 2006; General Medical Council, 2021). It underscores the importance of designing inclusive academic support structures that accommodate diverse educational backgrounds.

As shown in Figure 2, while some curricula were represented by smaller numbers, they may reflect distinct regional or institutional preferences. Overall, the distribution illustrates a student body heavily skewed toward internationally recognized curricula, particularly the American and British systems, while also representing a broad range of other educational frameworks from around the world. This diversity highlights the need for culturally responsive teaching and tailored pedagogical strategies to support equitable learning outcomes (Gay, 2018).

### Utility of Predictive Modelling in Medical Education

In applying these predictors, Bayesian Networks, Logistic Regression, and Neural Networks all yielded high-performing models, with AUC scores ranging from 0.91 to 0.94. These results affirm the value of predictive analytics in medical education, particularly in supporting early identification of students at risk of non-completion. Such tools could complement academic advising, enabling institutions like MBRU to provide more personalized academic support. This approach aligns with the findings of Kumar et al. (2021), who demonstrated that predictive analytics can accurately identify at-risk students in health professions education, enabling timely interventions to enhance student outcomes.

### Trends in Student Enrolment

As illustrated in Figure 1, the data suggest a positive trajectory in student enrollment over the years, with particularly strong growth in the most recent cohort. This increase may reflect the growing reputation of the medical program, expanded institutional capacity, or greater demand for medical education in the region. This trend aligns with national strategies aimed at advancing healthcare and expanding higher education, as highlighted in the UAE’s National Strategy for Higher Education 2030 (UAE Ministry of Education, 2030).

### Study limitations

Despite promising results, this study has several limitations. The analysis was conducted at a single institution, and model validation was performed internally. While internal validation results are encouraging, future research should include external validation using larger or multi-institutional datasets to assess the model’s generalizability beyond MBRU. Broader validation using data from other medical schools is necessary to confirm the generalizability of these findings. Additionally, the study focused on academic predictors and did not account for non-academic variables such as motivation, well-being, or socioeconomic factors, which could enrich future research. The study also focused on graduation as the primary outcome; however, it does not account for post-graduation success indicators, such as residency placement, licensing exam performance, or long-term professional achievement, which may provide a more comprehensive view of educational effectiveness.

## CONCLUSION

The findings of this study suggest that medical schools should consider placing greater emphasis on academic performance during the program, rather than relying solely on pre-admission criteria such as high school curriculum, to identify students most likely to succeed. While high school curriculum alone is not a strong standalone predictor of medical school graduation, its integration with academic indicators, particularly cumulative GPA, yields accurate and actionable predictive models. Moreover, the results underscore the importance of providing targeted academic support and early interventions, especially for students from diverse educational backgrounds. Future research should explore the inclusion of non-academic variables and leverage more advanced predictive modelling techniques to further refine our understanding of factors influencing student success. By embracing data-driven approaches, medical schools can design more effective admission processes and support systems that enhance student retention and success within increasingly diverse academic environments.

## Data Availability

Data are available from the Mohammed Bin Rashid University of Medicine and Health Sciences (MBRU) Institutional Data Access / Ethics Committee (contact via IRB) for researchers who meet the criteria for access to confidential data.

## Conflict of Interest

The authors declare that the research was conducted in the absence of any commercial or financial relationships that could be construed as a potential conflict of interest.

## Author Contributions

JM, project administration, conceptualization, methodology, formal analysis, data curation; writing, review, and editing.

AHK, methodology, formal analysis, writing-original draft, writing-review and editing, supervision, project administration.

LA, methodology, formal analysis, writing-original draft, writing-review and editing, supervision, project administration.

AJA, methodology, formal analysis, writing-original draft, writing-review and editing, supervision, project administration, funding acquisition.

## Funding statement

The author(s) declare that financial support was received for the research, authorship, and/or publication of this article. The authors would like to thank Mohammed Bin Rashid University of Medicine and Health Sciences (MBRU) for payment of the article processing charges.

## Acknowledgments

Fawad Hussain, Associate Professor of Artificial Intelligence, School of Computer Science, University of Birmingham (Dubai)

Kashif Rajpoot, Professor of Medical AI | University of Birmingham Dubai, Deputy Head of School of Computer Science (for Dubai) | University of Birmingham

## Notes

### Competing Interest Statement

The authors have declared no competing interest.

### Funding Statement

The author(s) received no specific funding for this work.

